# Social patterning and stability of intention to accept a COVID-19 vaccine in Scotland: Will those most at risk accept a vaccine?

**DOI:** 10.1101/2020.11.19.20234682

**Authors:** Lynn Williams, Paul Flowers, Julie McLeod, David Young, Lesley Rollins, CATALYST project team

## Abstract

Vaccination is central to controlling COVID-19. Its success relies on having safe and effective vaccines and also on high levels of uptake by the public over time. Addressing questions of population-level acceptability, stability of acceptance and sub-population variation in acceptability are imperative. Using a prospective design, a repeated measures two-wave online survey was conducted to assess key sociodemographic variables and intention to accept a COVID-19 vaccine. The first survey (time 1) was completed by 3436 people during the period of national lockdown in Scotland and the second survey (n = 2016) was completed two months later (time 2) when restrictions had been eased. At time one, 74% reported being willing to receive a COVID-19 vaccine. Logistic regression analyses showed that there were clear sociodemographic differences in intention to accept a vaccine for COVID-19 with intention being higher in participants of white ethnicity in comparison to Black, Asian, and minority ethnic (BAME) groups, and in those with higher income levels and higher education levels. Intention was also higher in those who were ‘shielding’ due to underlying medical conditions. Our results suggest that future interventions such as mass media and social marketing need to be targeted to a range of sub-populations and diverse communities.

## 1. Introduction

Vaccination will be vitally important in controlling future waves of the COVID-19 pandemic. Despite uncertainty regarding the specifics of many of the potential vaccines (e.g. efficacy and required doses), it is clear that high levels of overall public acceptance will be required. In recent years, vaccination rates have fallen and public confidence in vaccines is inconsistent [1-4]. The term ‘vaccine hesitancy’ refers to the ‘delay in acceptance or refusal of vaccines despite availability of vaccine services’ [5, 6]. The reasons for vaccine hesitancy are multi-levelled and complex, involving psychological, social and contextual factors [7-9]. Vaccine hesitancy was evident during the H1NI pandemic, which saw variable vaccine uptake, with non-uptake related to concerns about vaccine safety and perceptions of threat and risk [10-14]. Preliminary evidence from the current pandemic suggests that a sizable proportion of the public are currently either undecided or unwilling to receive a future vaccine for COVID-19 [15-20]. The importance of high levels of uptake was demonstrated in a recent study which suggests that in order to “extinguish an ongoing epidemic”, the efficacy of a vaccine as the sole intervention needs to be at least 80% when uptake is at 75%. If uptake is lower than this then an even more efficacious vaccine would be needed [21].

A recent global survey from Ipsos MORI [18], conducted in July and August 2020, of 20,000 adults from 27 countries found that 74% of people would get a vaccine if it was available. However, only 37% strongly agree that they would want to get it, while 37% somewhat agree. Large variations in acceptance levels were also shown across countries, ranging from 97% in China, to 54% in Russia. The primary reason that people gave for not wanting to accept the vaccine was worry about the side effects, followed by concern about the effectiveness of a vaccine, and not feeling at risk of contracting COVID-19 [18].

Emerging research also suggests that particular sub-populations have lower acceptance intentions. In France, during late March 2020, 26% of the sample reported that they would not accept a vaccine. COVID-19 vaccination hesitancy was more prevalent in those with low income, young women, and among older adults aged 75+, and it was associated with political views [19]. In Australia, more positive views were reported in April shortly after the introduction of lockdown, with 4.9% stating they would not get the vaccine, and 9.4% stating indifference. Here, COVID-19 vaccination hesitancy was associated with lower education levels and health literacy, and the belief that the threat of COVID-19 had been exaggerated [20]. A study conducted two months later in Australia, when restrictions had been eased, found that those who were unwilling or unsure about accepting a COVID-19 vaccine had increased by 10% [16], suggesting that COVID-19 vaccination levels may fluctuate depending on the context of infection rates and restrictions. A further study from the US reports an acceptance level of 67% and found that acceptance levels were lower in unemployed participants and in Black American participants [17].

Together, these initial studies suggest that COVID-19 vaccine acceptance differs both across countries and in different sub-populations within countries. The research to-date has been based on cross-sectional designs and has not yet explored how intention to accept a COVID-19 vaccine may change in line with rates of infection or lockdown restrictions within the same sample. We explored this question of intention to vaccinate against COVID-19 in a prospective survey in Scotland at two time points (during national lockdown and during the easing of restrictions). The present study has three key research questions (RQs):

RQ1: What proportion of people would accept a vaccine for COVID-19?

RQ2: Is COVID-19 vaccine acceptance stable over time in the context of different infection levels and restrictions?

RQ3: What socio-demographic factors are associated with intention to accept a future vaccine for COVID-19?

## 2. Materials and Methods

The present study consisted of an online survey at two time points. The first survey (time one) was conducted during lockdown restrictions (20th May-12th June 2020, weeks 9-12 of national lockdown). The second survey (time two) was conducted two months later (during August 2020) when lockdown restrictions in Scotland had been eased and there was little community transmission. Ethical approval was received from the University of Strathclyde Ethics Committee (Ref: 61/05/05/2020/A Williams) prior to commencement of the study. The data reported here are part of the larger CATALYST project which examined the changes that people had experienced during lockdown.

### 2.1. Participants and procedure

For time one, participants were recruited with convenience sampling through advertisements on social media, including Facebook and Twitter, which directed participants to Qualtrics where they could access the online questionnaire. Included in the survey for time one was an information sheet and consent form which participants read and signed before completing the survey. For time two, participants were re-contacted via email and provided with a Qualtrics link to the online questionnaire, inviting them to take part at time two.

### 2.2. Questionnaire

#### 2.2.1. Demographics and Health

Participants provided self-reported sociodemographic and health data at time one for the following variables: age (18-49, 50+); gender (female, male); ethnicity (white, BAME); annual household income (<£16,000, £16,000 – £29,999, £30,000 - £59,999, £60,000+); level of education (no qualifications/left school at 16, high school/college, university) and risk group/shielding status (i.e. those who had been classified as high risk based on underlying health conditions or age) (yes/no).

#### 2.2.2. COVID-19 vaccination intention

Intention to be vaccinated for COVID-19 was measured by self-report at time one and again at time two. Participants were asked “If a vaccine for Coronavirus (Covid-19) becomes available, would you want to receive it?” and provided response options “I *definitely* would not want to receive it”, “I *probably* would not want to receive it”, “Unsure”, “I *probably* would want to receive it” and “I *definitely* would want to receive it”.

### 2.3. Statistical Analysis

The COVID-19 vaccination intention response options of “I definitely would not want to receive it”, “I probably would not want to receive it”, and “Unsure” were coded as “Vaccine Hesitant” and the options “I probably would want to receive it” and “I definitely would want to receive it” were coded as “Vaccine Willing” to create dichotomous “willing” versus “hesitant” variables. Frequency responses to the vaccination intention questions were calculated (RQ1). In addition, a McNemar’s test was used to examine any changes in vaccine willingness across the two time points from lockdown (time 1) to the easing of restrictions (time 2) for those participants who completed the survey at both time points (RQ2). Univariate and multivariate logistic regression analyses were carried out to examine the socio-demographic factors associated with COVID-19 vaccination intention at time 1 (we selected time 1 intention as the dependent variable in order to maximise sample size) (RQ3). Due to issues relating to the representativeness of our sample and specifically the under-representation of males and those with lower levels of educational attainment (as shown in Table 1), we carried out post-stratification weighting of these variables based on the Scottish census data to correct for these imbalances. All of the analyses that we report are based on this weighed data in order to allow for greater extrapolation of the results to the Scottish population. All analyses were conducted using IBM SPSS Statistics (version 25) at 5% significant levels.

**Table 1.**
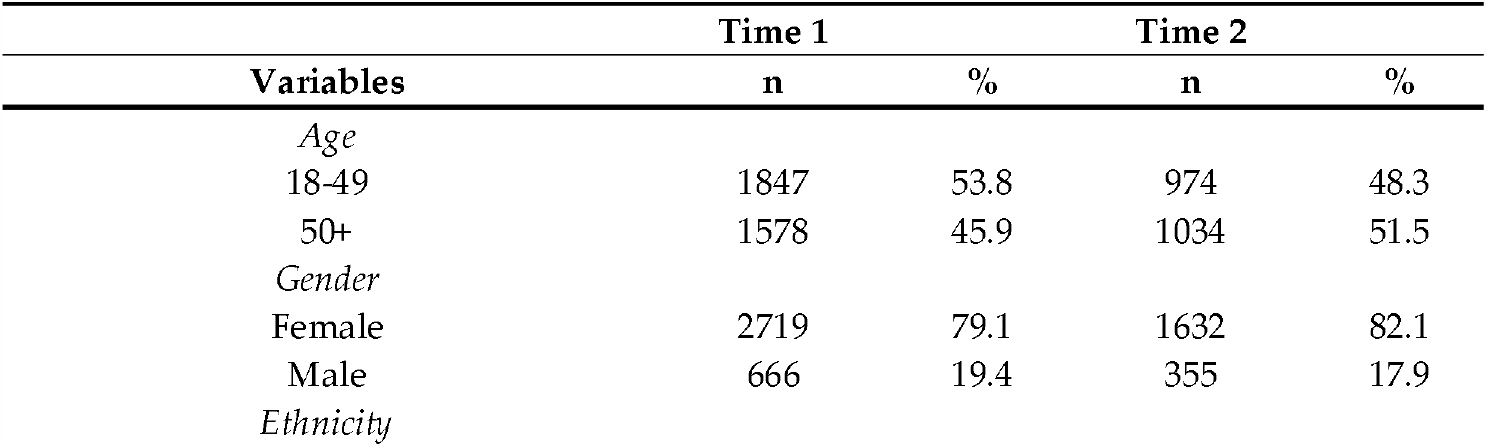
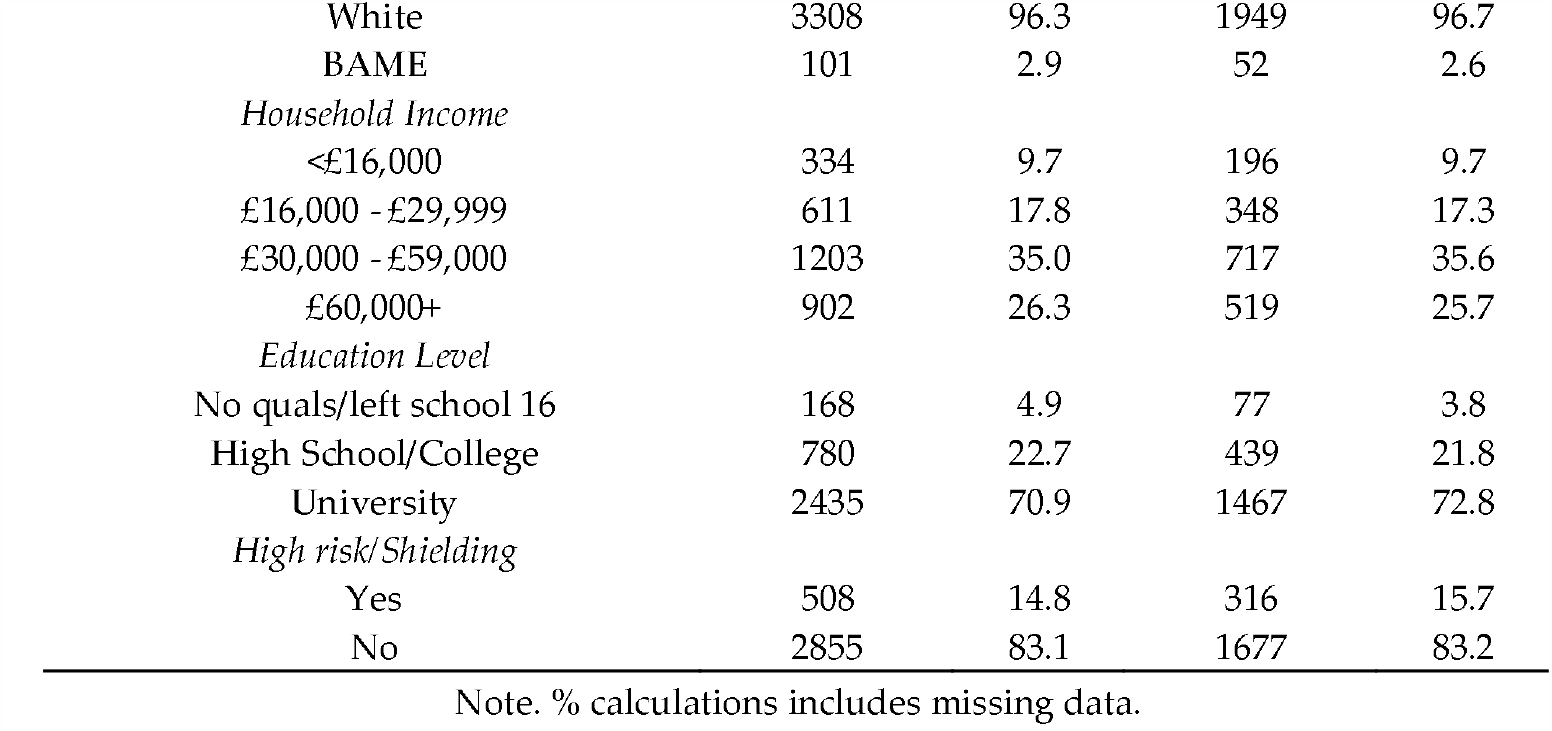
Sociodemographic and health variables for the sample at time 1 and time 2.

## 3. Results

Overall, 3436 participants (79% female) residing in Scotland, aged over 18, were recruited for the current study. The sample was comprised of participants aged between 18 and 92 (*M* = 46.21, *SD* = 15.26). Of the original sample, 2016 participants took part at time 2, representing a 59% follow-up rate. We compared the socio-demographic characteristics of those who participated in both waves of the survey with those who dropped out. There were no significant differences between the completers are non-completers based on ethnicity, income, or high risk status but the groups did differ significantly on education level, gender, and age with higher dropout in those with lower education levels, males, and in the younger age group. Participant socio-demographic characteristics from time one and time two from our original sample (not weighted) are shown in Table 1.

### 3.1. RQ1: What proportion of people would accept a vaccine for COVID-19?

A frequency analysis of the proportion of participants that reported willingness and hesitancy to accept a COVID-19 vaccine across the two time points are shown in Table 2. At time 1, 74% of participants were willing to receive a COVID-19 vaccine, and at time 2 this figure was slightly higher at 78% (note that the numbers reported in Table 2 for the ‘vaccine hesitant’ and ‘vaccine willing’ rows are based on the total sample, whereas the other numbers in the table are based upon only those participants who completed the questionnaire at both time points).

**Table 2.** Intention to accept a COVID-19 vaccine at time 1 and time 2.

| COVID-19 vaccine intention | Time 1 (national lockdown) |  | Time 2 (easing of restrictions) |  |
| --- | --- | --- | --- | --- |
|  | N | % | N | % |
| I definitely would not want to receive it | 54 | 3% | 65 | 4% |
| I probably would not want to receive it | 79 | 4% | 84 | 5% |
| Unsure | 301 | 17% | 262 | 14% |
| I probably would want to receive it | 498 | 27% | 506 | 27% |
| I definitely would want to receive it | 904 | 49% | 919 | 50% |
| Vaccine Hesitant | 850 | 26% | 416 | 22.5% |
| Vaccine Willing | 2406 | 74% | 1433 | 77.5% |

### 3.2. RQ2 – Is COVID-19 vaccine acceptance stable over time in the context of different infection levels and restrictions?

Table 2 presents the results relating to stability of COVID-19 vaccination intention for those participants who completed the questionnaire at both time points. McNemar’s test on these participants showed that there had been a significant shift in vaccine intention over time, (*p*=.004). Of the 54 participants who reported that they definitely would not want vaccinated at time 1, 13 (24.1%) reported that they would now accept the vaccine at time 2. Of the 79 who reported that they probably would not want the vaccine at time 1, 12 of them (15.2%) would now accept the vaccine at time 2. Of the 301 who were unsure, 113 (37.5%) reported at time 2 that they would now accept the vaccine. Of the 904 people who initially would definitely want to be vaccinated, 20 (2.2%) would now not take it, and 10 (1.1%) were now unsure. In addition, of the 498 who initially probably would receive it, 13 would now not take it (2.6%) and 72 are now unsure (14.5%).

### 3.3. RQ3: What socio-demographic and health factors are associated with intention to accept a future vaccine for COVID-19?

As shown in Table 3, univariate logistic regression analyses showed that there was a significant effect of age, ethnicity, education level, household income and high risk/shielding status on intention to accept a COVID-19 vaccine. There was no effect of gender on intention to accept a COVID-19 vaccine. In relation to age, younger participants reported higher levels of intention than those in the older age group. We also found that participants of white ethnicity had higher levels of intention than those from BAME groups. In relation to education, those with higher levels of education had higher levels of intention to receive the vaccine. In addition, those with higher levels of annual household income had higher intention levels than those with lower income levels. Finally, those participants at high risk or in the shielding category had higher levels of intention than those not in this group.

**Table 3.** Univariate analysis of socio-demographic factors and intention to accept a COVID-19 vaccine.

| Variable | p-value | Comparison | Coefficient | p-value |
| --- | --- | --- | --- | --- |
| Age | <0.001 | 50+ vs. 18-49 | 0.70 | - |
| Gender | 0.190 | Male vs. Female | 1.11 | - |
| Ethnicity | 0.018 | White vs. BAME | 1.72 | - |
| Education | <0.001 | High school/College vs. No qualifications/left at 16 | 1.98 | <0.001 |
|  |  | University vs. No qualifications/left at 16 | 2.78 | <0.001 |
| Household income | <0.001 | £16000 - £29999 vs. <£16000 | 1.11 | 0.441 |
|  |  | £30000 - £59999 vs. <£16000 | 1.39 | 0.009 |
|  |  | £60000+ vs. <£16000 | 1.97 | <0.001 |
| High risk/Shielding | 0.012 | Yes vs. No | 1.31 | - |

For the multivariate logistic regression we entered those variables that were significant in the univariate analysis (i.e. age, ethnicity, education, household income, and high risk/shielding). Ethnicity, education, household income and high risk remained significantly associated with intention to receive a COVID-19 vaccine in the multivariate analysis, but age was no longer significant. When considering the coefficients (see Table 4), those participants of white ethnicity are almost three times as likely to accept a COVID-19 vaccine compared to those from BAME groups. Similarly, those from the highest education group are two and a half times more likely than those from the lowest education group to accept it, and those in the highest income group are 1.82 times more likely to accept the vaccine compared to those in the lowest income group. In addition, those in a high risk/shielding group are almost twice as likely to accept a COVID-19 vaccine compared to those not in a high risk/shielding group.

**Table 4.** Multivariate analysis of socio-demographic factors and intention to accept a COVID-19 vaccine.

| Variable | p-value | Comparison | Coefficient | 95% CI | p-value |
| --- | --- | --- | --- | --- | --- |
| Ethnicity | <0.001 | White vs. BAME | 2.91 | 1.75-4.81 | - |
| Education | <0.001 | High school/College vs. No qualifications/left at 16 | 1.90 | 1.56-2.32 | <0.001 |
|  |  | University vs. No qualifications/left at 16 | 2.50 | 1.95-3.21 | <0.001 |
| Household income | <0.001 | £16000 - £29999 vs. <£16000 | 1.05 | 0.80-1.38 | 0.743 |
|  |  | £30000 - £59999 vs. <£16000 | 1.27 | 0.98-1.65 | 0.077 |
|  |  | £60000+ vs. <£16000 | 1.82 | 1.35-2.45 | <0.001 |
| High risk/Shielding | <0.001 | Yes vs. No | 1.95 | 1.53-2.49 | - |

## 4. Discussion

The present study is the first to examine intention to accept a COVID-19 vaccine in Scotland. The study also extends existing research that has examined acceptance levels in other countries by adopting a prospective design, allowing us to examine stability in vaccine intentions over time within the same sample. We found that 74% of the sample at time 1 intended to receive a COVID-19 vaccine (during the period of national lockdown). Interestingly, two-months later, intention levels had shifted significantly. The greatest shift in intention levels was apparent in the group who were “Unsure” at time one, with 38% of those participants now saying they would accept a vaccine. The second survey was carried out when restrictions had been eased and there was little transmission of COVID-19, it is therefore notable that intention to receive a COVID-19 vaccine was still high at this time. Stability in intention to receive a COVID-19 vaccination will be particularly important given that two doses of the vaccine are likely to be needed over time.

Our findings suggest that although the majority of respondents in Scotland indicated that they would want to receive a COVID-19 vaccine, there is a sizable minority of the public who were hesitant about receiving a vaccine, mirroring the overall picture that is emerging from other countries. Indeed, the intention levels reported here are similar to those emerging globally from the recent Ipsos survey in which 74% of people overall said they would get a COVID-19 vaccine [18].

We also found that intention to accept a COVID-19 vaccination varied by sub-population. There was a significant effect of ethnicity, education level, household income and high risk/shielding status on intention to accept a COVID-19 vaccine. Of particular note, those of white ethnicity are nearly three times as likely to accept a COVID-19 vaccine compared to those from BAME groups, those from the highest education group are two and a half times more likely than those from the lowest education group to accept it, and those in the highest income group are almost twice as likely to accept the vaccine compared to those in the lowest income group.

Overall, those most at risk of the negative sequalae of COVID-19 had less intentions to get vaccinated than those at lower risk [22]. For example, intention was higher in participants of white ethnicity in comparison to BAME groups. However, it should be noted that this binary approach to looking at differences in ethnicity is problematic, as it may mask important differences between diverse racialised and minoritised communities with distinct cultures and social norms. We also found that intentions for vaccination were higher in those with higher levels of income, again raising questions about why those most likely to suffer the negative consequences of COVID-19 would be the least likely to intend to vaccinate. These findings fit with the emerging literature on COVID-19 vaccine acceptance which suggests that those from BAME groups may be less likely to accept a vaccine [17] and that COVID-19 vaccine hesitancy is more prevalent in those with lower income [19]. If these inequalities in intention translate into uptake then these findings are very important as they suggest the likelihood that vaccination programmes, unless implemented with targeted and tailored interventions to enhance vaccination rates, may actually amplify existing inequalities. More positively, intention to accept a COVID-19 vaccine was higher among those who were shielding (i.e. those who had been classified as high risk based on underlying health conditions or age).

Together, these initial studies highlight the need for government and public health bodies to think carefully about how to approach their publics with further demands for behavioural change (i.e., uptake of a COVID-19 vaccine). The findings here suggest, for example, that a ‘one size fits all’ approach to mass media interventions represents at best a partial solution to increasing vaccination uptake and at worst, a solution that backfires, amplifying existing inequalities. These findings suggest that future interventions need to be targeted to a range of sub-populations and diverse communities [23, 24]. The range of interventions should offer different kinds of educational, persuasive or enabling approaches [25] to different groups of people and there is growing recognition that such interventions should be co-produced. Issues of visual cultural representation, identifiable local key opinion leaders, readability and reading levels, and trust will all be important considerations. These emerging findings also lend themselves to considering the wider opportunities to targeting afforded by social media. Whilst marketing and politics are capitalising on these new opportunities, it is less clear if and how public health can do so. Equally these initial findings also suggest that health care professionals should be trained to anticipate and respond to diverse segments of the population differently with overt and inclusive demonstrations of cultural competencies.

If we imagine a future of targeted interventions to improve COVID-19 vaccination uptake, it is also important to focus now on the granularity of messaging that will shortly be needed. Whilst evidence is emerging concerning where and amongst whom interventions need to be targeted, it is currently unclear how intervention content should be tailored to these populations and communities and their particular beliefs. We need to use the full range of social and behavioural sciences now to address this key gap and ensure that we are prepared and ready to support the whole population in gaining maximum benefit from available vaccines. One preliminary study in this area used the Behaviour Change Wheel [26-28] to provide recommendations for the design of interventions aimed at maximising public acceptance of COVID-19 vaccine. The findings suggested that interventions should utilise the behaviour change techniques [29] of information about health, emotional, social and environmental consequences, and salience of consequences in order to provide the public with information about the beneficial consequences for themselves and for others [30]. Further, more in-depth research like this which focuses on intervention development is required.

Strengths of the current study include the large sample size, the use of two waves of data collection to examine stability in COVID-19 vaccination intention and the inclusion of a range of sociodemographic factors to allow us to understand the social patterning of COVID-19 vaccination acceptance. However, it is important to note that the study is not nationally representative, and weighting was applied to the gender and education variables due to the under-representation of males and those with lower levels of education. Furthermore, we experienced a participant drop-out rate of 41% from time one to time two meaning that we cannot draw any conclusions about the change in COVID-19 vaccination intention in those participants who were lost at follow-up. Moreover, we experienced higher dropout in those with lower education levels, males, and in the younger age group. In addition, participants were answering about their intention to accept a hypothetical COVID-19 vaccine, without information regarding the specifics of the vaccine in terms of number of doses needed, potential side-effects, or prioritisation of delivery across the population. All of which may influence eventual uptake of a COVID-19 vaccination.

## 5. Conclusions

Intention to accept a COVID-19 vaccine is currently high in Scotland and our findings suggest that intention to receive the vaccine did not fall in the context of lower infection rates and fewer restrictions. However, the data also point to a sizable minority of the public who are hesitant about receiving a future COVID-19 vaccine. Of note, intention was higher in participants of white ethnicity in comparison to those from BAME groups, and in those with higher levels of income and education. Our findings and those from other studies suggest that future interventions need to be targeted to a range of sub-populations and diverse communities. To do so, we need to better understand the barriers to vaccination in these groups so that we can collectively be better prepared to deliver appropriate evidence-based culturally and community-appropriate messaging aimed at maximising COVID-19 vaccine uptake.

## Data Availability

Data will be made available upon publication of the paper.

## Author Contributions

Conceptualization, L.W. and P.F; methodology, L.W and L.R..; formal analysis, J.M and D.Y.; investigation, L.R; writing—original draft preparation, L.W., P.F., J.M.; writing—review and editing, all authors.; funding acquisition, L.W and P.F. All authors have read and agreed to the published version of the manuscript. The CATALYST project team: Leanne Fleming, Xanne Janssen, Alison Kirk, Madeleine Grealy, Bradley MacDonald.

